# Real-time time-series modelling for prediction of COVID-19 spread and intervention assessment

**DOI:** 10.1101/2020.04.24.20078923

**Authors:** Taha Hossein Rashidi, Siroos Shahriari, AKM Azad, Fatemeh Vafaee

## Abstract

Substantial amount of data about the COVID-19 pandemic is generated every day. Yet, data streaming, while considerably visualized, is not accompanied with advanced modelling techniques to provide real-time insights. This study introduces a unified platform which integrates visualization capabilities with advanced statistical methods for predicting the virus spread in the short run, using real-time data. The platform is backed up by advanced time series models to capture any possible non-linearity in the data which is enhanced by the capability of measuring the expected impact of preventive interventions such as social distancing and lockdowns. The platform enables lay users, and experts, to examine the data and develop several customized models with different restriction such as models developed for specific time window of the data. Our policy assessment of the case of Australia, shows that social distancing and travel ban restriction significantly affect the reduction of number of cases, as an effective policy.

## Introduction

The outbreak of coronavirus disease 2019 (COVID-19), caused by acute respiratory syndrome coronavirus 2 (SARS-CoV-2), has been recognized as a pandemic by World Health Organization representing the most serious public health threat during the last century [1]. The global impact of COVID-19 has been profound. As of 12 April 2020, more than 1.87 million cases of COVID-19 have been reported in over 200 countries, resulting in 118,851 deaths as reported by the European Centre for Disease Prevention and Control (ECDC) [2].

Forecasting the imminent spread of COVID-19 informs policymaking and enables an evidence-based allocation of medical resources, arrangement of production activities and economic development [3]. Therefore, it is urgent to establish efficient trend prediction models, on the latest available data, to provide a point of reference for the governments to formulate adaptive responses based on reliable predictions on the impending progress of the pandemic.

The classical Susceptible-[Exposed]-Infected-Recovered (SEIR/SIR) epidemic models [4], have been widely developed to simulate the transmission dynamics of COVID□19 [5, 6] and the impact of non-therapeutic interventions -e.g., travel and border restrictions [7, 8], quarantines and isolations [5, 9-11], or social distancing and closure of facilities-on the spread of the outbreak, and in some cases, on the healthcare demand [5, 9, 11-13].These studies have been mostly focused on calibrating models for a specific country/region based on the data at the time of the model-development and assuming a multitude of parameters initialized upon prior knowledge such as social contact structure, rate of compliance with the policy and incubation or infection period among others. Complementing upon SEIR mathematical models, and owing to the increased amount of data and consistency of reports, some recent efforts have been focused on developing statistical [3, 14] or machine learning methods [15] to predict the near-future spread of COVID-19 (in terms of the number of confirmed cases or deaths) based on the historical data.

While reliable predictions of the pandemic trend are essential for policymaking and resource-allocation, there is a lack of an adaptive real-time modelling platform which evolves as new data arrives. In response to this urgent need, we present an advanced time-series models for the progression of COVID-19 using the Autoregressive Integrated Moving Average (ARIMA) formulation [16] statistical analyses combined with several non-linear transformation approaches [17], and complemented with an interactive online dashboard which efficiently generates country-wise predictive models, in real-time, based on the latest ECDC report of COVID-19 cases worldwide.

The proposed modelling approach neither relies on strict modelling assumptions (e.g., linearity, stationarity, or existence of an epidemic steady state) nor on any initial parameters requiring a priori knowledge. It offers a transparent mathematical function to better understand the trend and to predict future points in the series. Different types of transformation have been examined to capture the nonlinearity in the time-series data followed by multiple differencing steps to eliminate the non-stationarity status. Notably, we enhanced the time-series model to capture the effect of previous interventions using an exogenous variable which can be used to predict the impact of future interventions. Further, when no record of intervention is provided, the model can infer previous interventions from data and incorporate its estimated impact into future predictions.

The main objective of this study is to introduce an easy-to-use and readily available statistical tool to develop rigor models for time series data of COVID-19 as data becomes available on a real time basis. In this article it is demonstrated that the proposed modelling tool is reliable to estimate accurate model parameters and is capable of being used for policy assessment. The tool is perfectly tailored for modelling COVID-19 data which an option of assessing the performance of any interventions to control the spread, such as lockdown, social distancing rules, and airport restrictions.

### Model Development and Performance

Multiple transformation operations are investigated to stabilise variance, coupled with recursive differencing until eliminating non-stationarity in the time-series data, i.e., p-value < 0.05 based on augmented Dickey–Fuller test [16]. Upon each transformation, the best ARIMA model is obtained for each country, according to Akaike information criterion (AIC) value using maximum likelihood estimation. The optimal model for each transformation is then recorded based on the overall model Root Mean Square Error (RMSE) on the last 20% of observations reported as a surrogate estimate of out-sample prediction performance. The predictive power of the best model per country is compared against estimations provided through 1) exponential growth in number of cases, 2) doubling time of two days, 3) doubling time of 3 days and 4) doubling time of one week, as well as a conventional linear univariate regression on log-transformed data. Table 1 shows the parameters of the optimal ARIMA model per country and the corresponding RMSE measures (of the last 20% of observations) compared with conventional trends (based on ECDC data on April 13, 2020). While, the purpose of this study is not to develop the most accurate time-series predictive model, statistics of Table 1 clearly show that using a more sophisticated statistical model significantly improves the prediction accuracy of COVID-19 spread in the near future (t-test p-value << 0.001 comparing residuals’ distributions), which signifies the urgency of such studies for policy appraisal. In other words, having access to tools, such as the one introduced in this study, enables experts with limited knowledge about details of statistical specifications to readily use such specifications to nowcast and forecast the effectiveness of policies they envision and propose for controlling the spread of COVID-19, or similar outbreaks.

**Table 1:** Model specifications and performance. Only countries with more than 35 days after the observation of the 50^th^ case are included.

| Country | Transform<br>ation | Model<br>ARIMA(p, i, q) | Parameters (SE) | RMSE (p-value vs ARIMA) |  |  |  |  |
| --- | --- | --- | --- | --- | --- | --- | --- | --- |
|  |  |  |  | ARIMA | Linear Regression | Doubled in 2 days | Doubled in 3 days | Doubled in 7days |
| Australia | Ratio | ARIMA(1,2,2) | AR1: -0.512 (0.18), MA1: -1.312 (0.19), MA2: 0.396 (0.2) | 50.76 | 5830.71 (1.55E-03) | 365042.98 (1.55E-03) | 2728.15 (1.55E-03) | 6043.16 (1.55E-03) |
| Austria | Without | ARIMA(4,5,0) | AR1: -0.393 (0.15), AR2: -0.118 (0.16), AR3: 0.082 (0.16), AR4: 0.402 (0.14) | 86.4 | 11429.25 (5.83E-04) | 128626.51 (5.83E-04) | 10665.14 (5.83E-04) | 12916.79 (5.83E-04) |
| Canada | Without | ARIMA(0,4,1) | MA1: -0.48 (0.13) | 133.38 | 11180.07 (5.83E-04) | 122299.91 (5.83E-04) | 17162.77 (5.83E-04) | 19510.36 (5.83E-04) |
| China | Ratio | ARIMA(0,1,1) | MA1: -0.726 (0.07) | 20.64 | 280692.59 (5.96E-11) | 2.51759E+14 (5.96E-11) | 3157347232 (5.96E-11) | 74300.1 (5.96E-11) |
| France | Ratio | ARIMA(0,1,2) | MA1: -1.328 (0.18), MA2: 0.441 (0.19), INTERCEPT: -0.009 (0) | 1329.8 | 97137.98 (1.75E-04) | 1907006.52 (1.75E-04) | 70032.2 (1.75E-04) | 82691.58 (1.75E-04) |
| Germany | Without | ARIMA(0,4,1) | MA1: -0.378 (0.18) | 960.29 | 152545.37 (4.11E-05) | 1886859.27 (4.11E-05) | 95897.65 (4.11E-05) | 108521.16 (4.11E-05) |
| Greece | Logaritmik | ARIMA(0,2,1) | MA1: -0.562 (0.2) | 20.89 | 642.31 (5.83E-04) | 97052.71 (5.83E-04) | 701.73 (5.83E-04) | 1872.59 (5.83E-04) |
| Iceland | Without | ARIMA(0,4,1) | MA1: -0.598 (0.11) | 8.28 | 647.89 (5.83E-04) | 97321.26 (1.75E-02) | 779.17 (1.75E-02) | 1585.79 (5.83E-04) |
| Iran | Without | ARIMA(0,4,2) | MA1: -0.909 (0.14), MA2: 0.354 (0.14) | 250.61 | 52502.63 (4.11E-05) | 5536118.99 (4.11E-05) | 38379.54 (4.11E-05) | 62048.14 (4.11E-05) |
| Iraq | Ratio | ARIMA(1,1,1) | AR1: -0.533 (0.15), MA1: -0.77 (0.13) | 73.44 | 187.33 (2.16E-03) | 36466.36 (2.16E-03) | 343.45 (2.16E-03) | 1168.51 (2.16E-03) |
| Italy | Logaritmik | ARIMA(0,2,3) | MA1: -0.812 (0.12), MA2: -0.194 (0.17), MA3: 0.62 (0.12) | 432.34 | 158551.98 (1.08E-05) | 10499769.78 (1.08E-05) | 98692.2 (1.08E-05) | 134369.1 (1.08E-05) |
| Japan | Ratio | ARIMA(4,0,0) | AR1: 0.507 (0.13), AR2: -0.455 (0.14), AR3: 0.491 (0.14), AR4: -0.416 (0.14), INTERCEPT: 1.09 (0.01) | 431.49 | 371.84 (2.84E-06) | 114429661.6 (2.84E-06) | 201685.67 (2.84E-06) | 3964.28 (2.84E-06) |
| Norway | Without | ARIMA(0,4,1) | MA1: -0.566 (0.13) | 51.71 | 25.86 | 4303.38 (1.55E-04) | 365023.96 (1.55E-04) | 2666.24 (1.55E-04) |
| Singapore | Ratio | ARIMA(0,1,1) | MA1: -0.749 (0.11) | 155.26 | 264.49 (7.40E-07) | 309923455 (7.40E-07) | 392790.3 (7.40E-07) | 1263.96 (7.40E-07) |
| South Korea | Ratio | ARIMA(3,1,0) | AR1: 0.164 (0.13), AR2: -0.358 (0.14), AR3: -0.578 (0.15), INTERCEPT: -0.013 (0.01) | 29.79 | 6466.12 (2.17E-05) | 29989620.31 (2.17E-05) | 76833.4 (2.17E-05) | 10214.61 (2.17E-05) |
| Spain | Ratio | ARIMA(0,1,3) | MA1: -0.65 (0.14), MA2: -0.256 (0.16), MA3: 0.362 (0.16) | 763.79 | 200633.37 (3.11E-04) | 927627.11 (3.11E-04) | 135512.96 (3.11E-04) | 144107.03 (3.11E-04) |
| Sweden | Ratio | ARIMA(0,1,0) |  | 157.69 | 2516.42 (1.17E-03) | 189750.17 (1.17E-03) | 5588.52 (1.17E-03) | 8497.51 (1.17E-03) |
| Switzerland | Without | ARIMA(1,4,0) | AR1: -0.485 (0.14) | 223 | 23676.88 (1.55E-04) | 350861.71 (1.55E-04) | 18990.29 (1.55E-04) | 23163.8 (1.55E-04) |
| UK | Ratio | ARIMA(0,1,1) | MA1: -0.667 (0.15) | 2222.5 | 35552.09 (1.55E-04) | 317174.36 (1.55E-04) | 55564.92 (1.55E-04) | 60024.61 (1.55E-04) |
| USA | Without | ARIMA(1,5,3) | AR1: 0.914 (0.08), MA1: -1.398 (0.25), MA2: 0.275 (0.22), MA3: 0.363 (0.15) | 3525.5 | 471365.84 (4.11E-05) | 5231573.73 (4.11E-05) | 384278.2 (4.11E-05) | 410846 (2.88E-04) |

**Abbreviations:** **ARIMA:** Auto Regressive Integrated Moving Average, **SE:** Standard Error, **RMSE:** Root Mean Square Error, **AR:** Autoregression coefficients, **MA:** Moving average coefficients, **(p, i, q):** *p* the order, i.e., number of time lags, *i* the degree of differencing, and *q* is the order of the moving-average model

### Effect of Transformation

Different time-series transformation operations, namely power transformation, logarithmic transformation and ratio transformation, have been applied to pre-process the data prior to the differencing step. We have observed that the type of transformation can significantly improve the performance of a model (in terms of the estimated out-sample RSME) as there is no a *priori* knowledge about the best-performing transformation (except that power transformation always performs poorly). Figure 1A shows some countries, as case studies, whose ARIMA models (as of April 13, 2020) are significantly affected by the type of transformation. As Figure 1A shows some countries such as Canada has significantly better performance (t-test p-value < 0.05) without any transformation. Italy’s model performs better (p-value < 0.06) by using ratio or logarithmic transformations and for USA, transformations do not significantly affect the performance (p-value > 0.6 for all comparisons). The case Greece, quite interestingly, shows that the ratio transformation stands above the other two while there is no statistically significant evidence that the logarithmic transportation performs any better than the no transformation case. Overall, the results signify the value of a performance-driven transformation selection approach upon trying multiple operations, as implemented in this platform.

**Figure 1.**
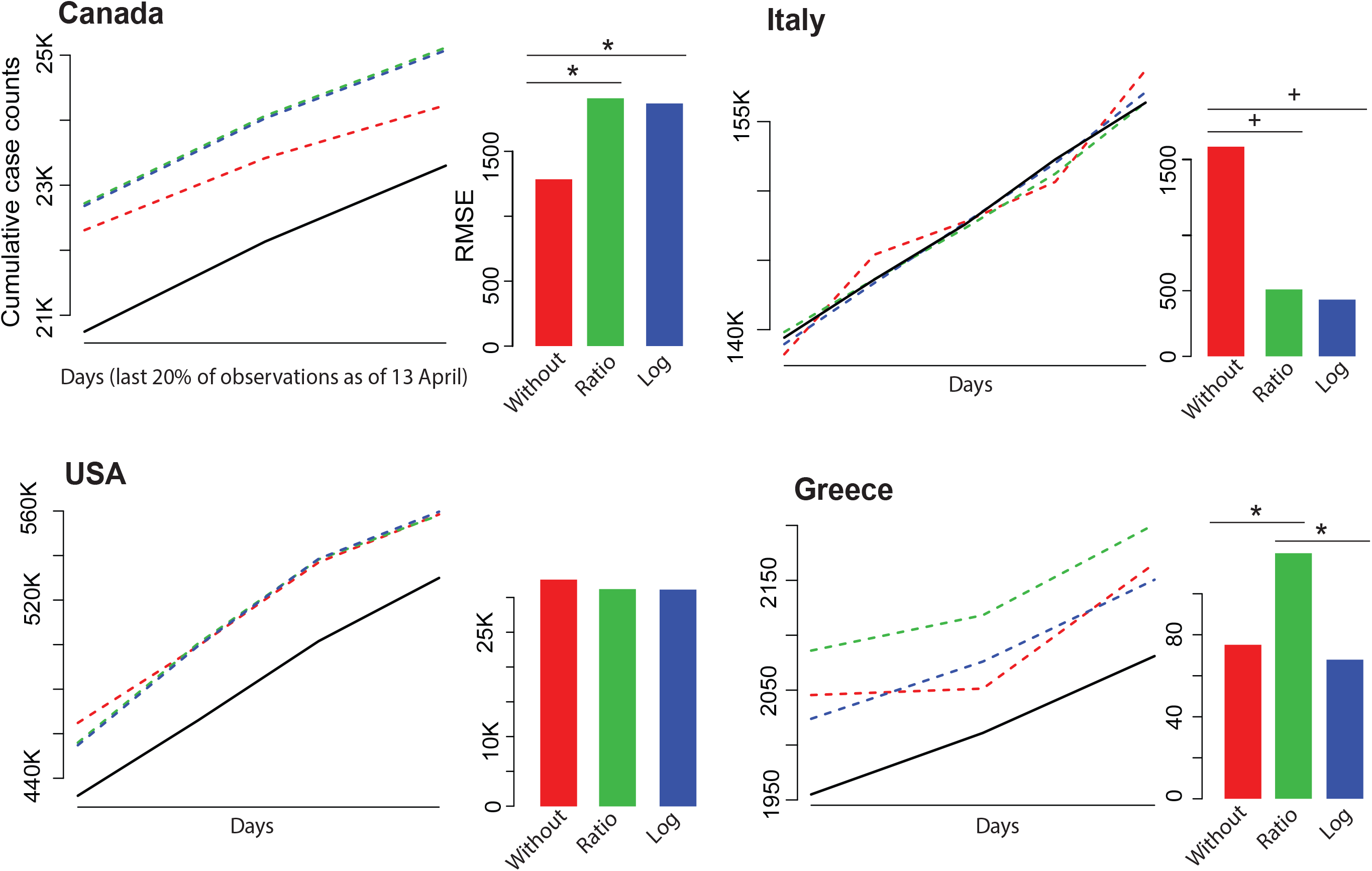
Effect of transformation on modelling performance. Four countries were selected as case studies to demonstrate the effect of ratio and logarithmic transformations on the model performance as measured by RMSE on last (most recent) 20% of time-series data. The solid line shows the observed trend and the dashed lines shows model fitted values without transformation (red) and after ratio (green) or logarithmic (blue) transformation. The bar plot beside each trend graph shows the corresponding RMSE estimations. ‘*’ implies that t-test p-value < 0.05 while ‘+’ implies that p-value < 0.06.

**Figure 2.**
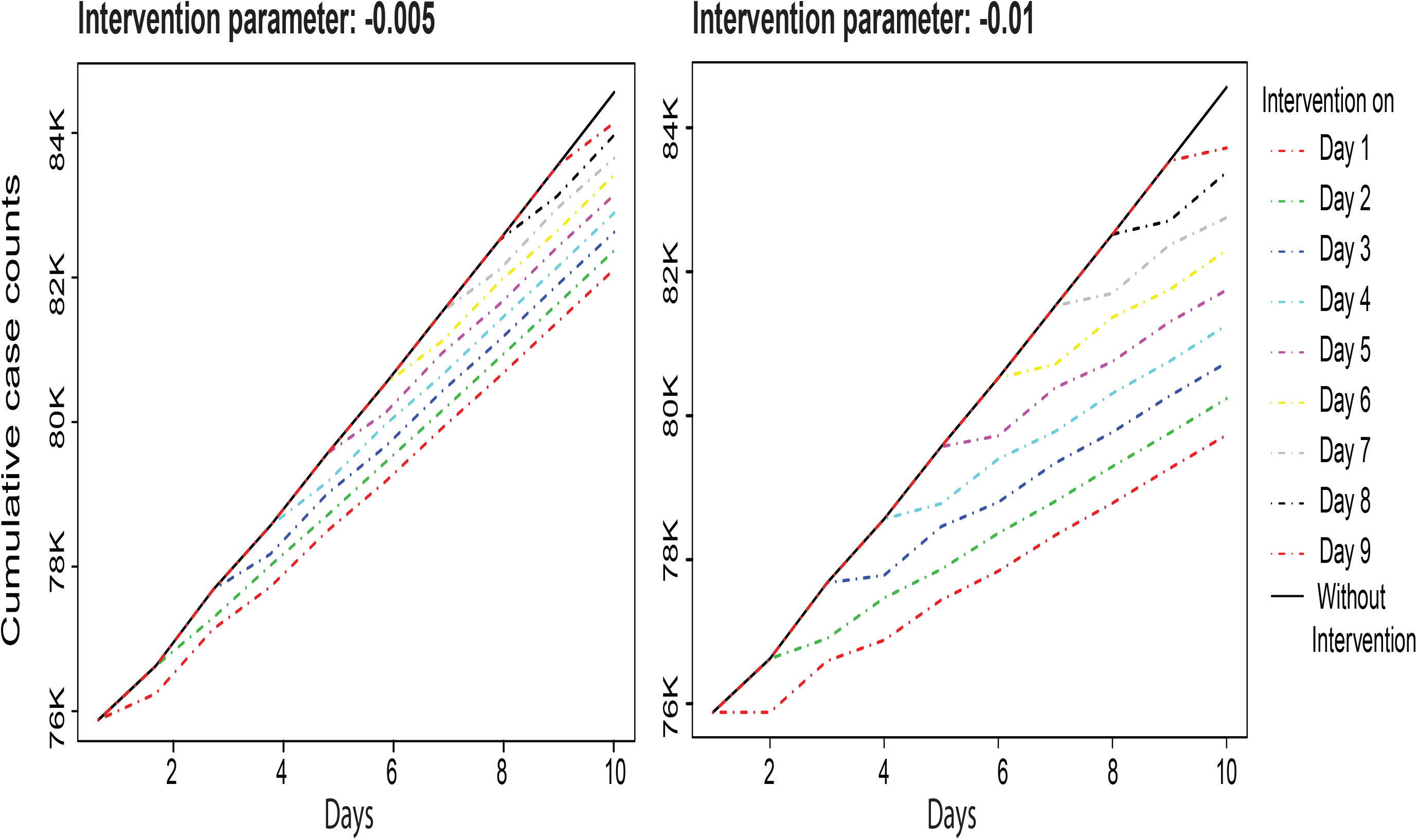
Sensitivity analysis on the impact of the intervention parameter. Left if the model with the intervention parameter of -0.03, and right is the model with intervention parameter of -0.01. Both models are based on the results obtained for the China data.

### Dynamic Model Estimation

The nonlinear dynamic system underlying COVID-19 spread is producing a regularly disrupted pattern making static predictions increasingly unreliable. Accordingly, a powerful feature of the platform is *dynamic model estimation*, that is, all models are re-optimised temporally with availability of new daily observations. Accordingly, the latest reports on COVID-19 case numbers are reflected in model estimation which accounts for the impact of new interventions improving the reliability of the future forecasts. As a case study, we have chosen to show the value of this feature on prediction of future case numbers in Iran. Iran’s trend shows significant fluctuations in the last 10 days (as of April 13, 2020) offering an interesting case study. We assumed that the model has access to data up to April 03, and then reported the next 10 days predictions and the RMSE of predicted number on April 13th. This procedure was repeated 10 times, where new observations became available to the model, one at the time. Figure 3 shows how such dynamic re-estimation adjusts the model with emerging pattern in time-series trend and improves prediction accuracy.

### Intervention Detection

The modelling platform developed in this study can identify unreported interventions to adjust the estimated models based on any exogenous fluctuations in the data. In other words, assume that some fluctuations occurred in the time-series (for instance, due to quarantines, changes in the reporting or testing methods, or facility closures), for which are reliable reported information is not available. These fluctuations can be inferred by using the platform to add the inferred interventions as an external regressor to the model to correct the bias in the parameter estimation which will be used later for prediction purposes.

The procedure for detecting intervention is to iteratively add interventions on potential dates, re-estimating the model and monitoring changes in the specification of the ARIMA model. As soon as major changes are observed in the model structure, i.e., values of (p, i, q), such alterations can be interpreted as an intervention in the data and the identified days would be associated with the dates of inferred interventions. The external regressor, corresponding to the interventions, is basically a binary variable which, for each day, is 0 (default) indicating no intervention and 1, otherwise. The coefficient and the significance of the intervention variable reflect the average impact of the recorded interventions. The online platform, as it is further discussed in the next subsection, enables users to incorporate the effect of known interventions into model specifications, and to detect unknown potential interventions which can capture observed fluctuations in the disease spread time-series.

The proposed intervention modelling approach is examined on the data provided for China as reported dates of interventions are officially available. To examine the effectiveness of the proposed intervention detection procedure, three models are developed, on the number of cases reported from the end of December to the 20^th^ of February, with 1) no intervention, 2) reported interventions, and 3) inferred interventions. The model with no interventions returns the RMSE of 6.52, the one with reported interventions returns the RMSE of 6.53 and the one with the inferred interventions returns the RMSE of 6.40. In the reported intervention model, three days are obtained from the available information online about major interventions in China happening on 23th, 24th, and 27th of January 2020, while the inference approach identified the following intervention days as 20th, 24th, 26th, 29th, 30th of January 2020, which are generally lagged compared to the reported ones, possibly because the impact of these quarantine decisions are reflected in the system with a delay. The proposed intervention inference approach, that automates identification of the interventions, finds a statistically significance coefficient for the average impact of the inferred interventions and can successfully improve the goodness-of-fit of the model.

The estimated coefficient of -0.04 (which is statistically significant at the 80% confidence level unlike the parameter of the model with reported interventions in the inferred intervention model has a major impact on reduction of number of cases (almost no increase) if used to predict the number of cases to the stable situation of China. Therefore, we performed some sensitivity analysis on the impact of this variable on the prediction results which are presented in Figure 2. The left graph shows the impact of an intervention applied on different days in the next 10 days where the intervention parameter is set to -0.005 and the right graphs shows the situation that a coefficient of -0.01 is considered. The main take away message of the diagrams of Figure 2 is that it is better to apply the interventions as early as possible even if the impact of the intervention is as small as the one considered in the right graph. Further, stronger interventions, not only have a larger immediate impact, but they result in a more stable long-term impact as shown in Figure 2.

**Figure 2.**
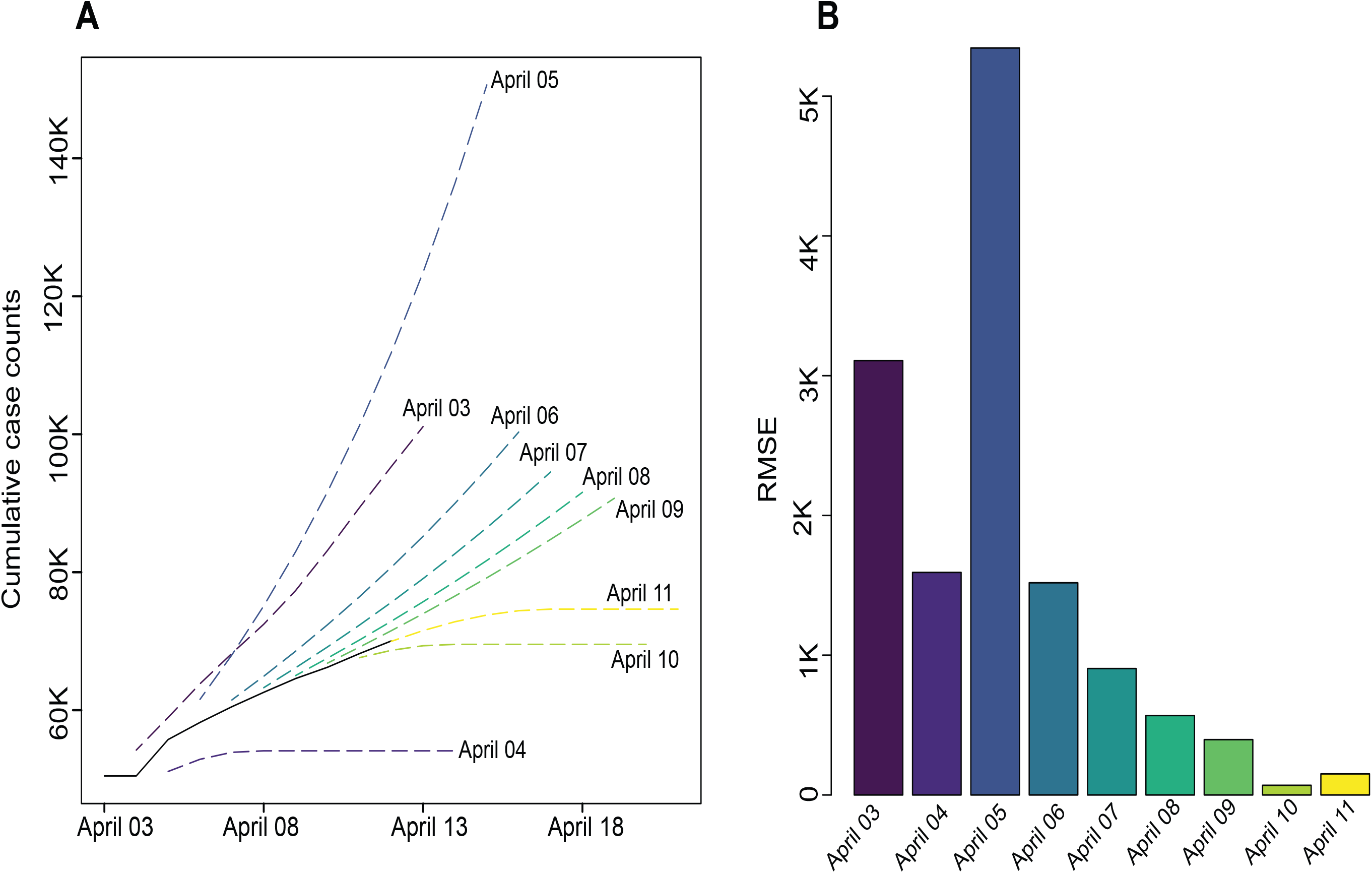
Dynamic Model Estimation. **A**. Predictions of next 10 days for COVID trend in Iran, assuming that the last available date was April 03 2020 to April 11 2020 (last obsereved date at the time of analysis: April 13). **B**. RMSE comparing predictions with obsereved data at April 13.

### Assessing the Effectiveness of Containment Policies (in Australia)

To further assess the effectiveness of policies using the proposed platform, we have looked at the response of the Australian government to COVID-19, given the access of the authors to detail information on timeline of interventions by the Australian government. The first case in Australia was confirmed on January 25^th^ by a passenger travelling from Wuhan which was followed by a travel ban from mainland China on February 1^st^. Other travel bans were followed by, Iran arrival blocking on March 1^st^, south Korea on March 5^th^, Italy on March 11^th^, self-isolation for overseas travelers and cruise ships blocked on the 15^th^ of March, border closure on March 19^th^, ban of travelling overseas on 24^th^, and mandatory isolation in hotels for travelers on 28^th^ [18]. Upon including travel bans in the model (data from 5^th^ March to 20^th^ of April), the estimated coefficient of the intervention variable would be -53.89 (with standard error, SE = 26.94) implying that on average the travel bans could reduce the number of infected people by 53 cases. The model predictive power would be significantly improved in terms of the goodness of fit of the model (RMSE = 8.88, *significant* compared to ‘no intervention’ residuals). Other than the coefficient of the travel ban intervention, an ARIMA (1, 1, 2) is estimated reflecting the importance of incorporating the observation of three days before into prediction, which also requires one differencing to make the data stationary reflecting the non-linearity of time-series data. The first order of integration (*i*=3 in the ARIMA model) also implies that the slope of the number daily cases is critical in the model, not just the accumulated total number of reported cases.

Another type of intervention practiced in Australia has been restrictions on gathering in public places. Australia imposed restrictions on outdoor gathering of people to less than 500 on the 16^th^ of March, indoor gathering to less than 100 individuals on 18^th^ of March, Pubs clubs closed, restaurants take-away only on 23^rd^ of March, and all gathering limited to 2 persons on 30^th^ of March [18]. By considering these interventions except for the 23^rd^ which over laps with a major spike in the data, the parameter of intervention variable is estimated as -16.24 (SE = 38.46) imply that the gathering bans in Australia reduced the number of cases by -16 cases per day, while the coefficient is not significant. The corresponding ARIMA models is (3,3,0), with RMSE 10.31 (*not significant* compared to ‘no intervention’ residuals).

Once all restrictions are included in one model to examine the effect of interventions in one model with the aim of finding the best fit to the data (on 6^th^, 12^th^, 15^th^, 16^th^, 19^th^, 24^th^, and 28^th^ of March and 3^st^ of April) an ARIMA (1,1,2) model is obtained with RMSE of 8.68, where the coefficient of intervention variable is -45.4 (SE=21.86, p-value = 0.03) and the model is only marginally improved to the first model. In other words, the intervention strategies of the government of Australia could result in a reduction of 45 cases per day during March and early-to-mid April 2020. As a result, by considering the first model and the last model which is developed to find the best fit to the data we can say that the preventive strategies helped reducing the number pf infected cases in Australia by 45-53 cases.

### Online Dashboard

We have developed an interactive online dashboard (https://unsw-data-analytics.shinyapps.io/covid19_analytics) to facilitate real-time model development for lay users as well as data scientists. Users can select the country of interest from the left panel and observe an interactive visualisation on cumulative counts of confirmed cases in the middle panel. Upon pressing the ‘Predict’ button’, the platform provides users with optimal models fitted to the latest reports of COVID-19 spared as provided by ECDC. For any country of interest, the interactive user-interface enables users to re-estimate models by 1) adding observed interventions in previous days, 2) customising the range of days to be included in the model and 3) incorporating the effect of future interventions in predictions. The right panel visualises the cumulative number of confirmed cases since the 1000^th^ case of top 10 countries in terms of total number of cases, plus predictions of growth trajectories in the next 10 days. Similarly, the middle-button panel shows the world map color-coded with predicted number of cases per 100K, together providing a global comparative view of the forthcoming COVID-19 spread.

## Discussion and Conclusion

Real-time COVID-19 data analytics have been mainly focused on visualizing the spread [19] with limited effort in developing models to dynamically analyze the data. Epidemiological models, i.e., SIR/SIER models, have a strong foundation in analyzing epidemic growth/decline, and have been substantially explored for modelling the *speed* of infectious disease progression. Yet, such models are often offline/static, require assumptions for the parametric formulation of the model and rely on multitude of initial parameters.

We developed a time-series based statistical model to dynamically predict the future trend of COVD-19 spread. It is built upon the strength of SIR/SIER models in considering the *speed* of progression and coupled with the capacity of time-series models in 1) considering higher orders of derivatives of the number of cases in previous time intervals, 2) accounting for the impact of residuals of the previous time intervals, and 3) incorporating intervention effect as external variables. We presented an automated modelling platform that delves into multiple layers of information in the COVID-19 time series data to find the best fit with the aim of providing robust forecasts. The platform—unlike numerous counterparts that are primarily limited to disseminating the existing patterns—measures the average impact of previous interventions to reduce the growth of the infected cases. This measure is then used for predicting the future spread when similar interventions are considered to further control the outbreak.

The presented platform was shown to be effective in estimating the trend of outbreak for each country. We elaborated the importance of data transformation as a preprocessing step and shown that there is no transformation operation which consistently provides the best fit to the data. Hence, exploring multiple options are recommended to stabilize variations prior to modelling using conventional econometrics formulations. A unique aspect of the presented platform is that it facilitates real-time model development incorporating latest reported data into modelling. We have shown that such adaptive model estimation significantly improves the prediction power and therefor, forecasting reliability.

One major challenge ahead of governments and policymakers, is the uncertainty around the effectiveness of containment policies in controlling the outbreak. Yet, authorities can observe the effectiveness of what they have done in the past few days, weeks, and months and learn from the impact of their previous decisions to enhance their intelligence in proposing new mechanisms to control the outbreak. We have shown that examining the effect of controlling policies can inform policymakers on number and types of controlling policies required to achieve their objective in reducing the number of infected cases. We have shown for Australia, as a case study, that containment policies not only decline the number of infected cases right after implementation, but also reduce the slope of progression. The latter is a more outstanding factor, especially when such policies are coupled with economic related policies given the recession or depression that is expected to happen during or after the COVID-19 pandemic. Overall, given the unknown nature of SARS-CoV-2 spread, we need to relax the boundaries of potential methods, beyond classical epidemic models, to further explore the behavior of data to account for unknown aspects the virus spread.

## Data Availability

Provided in th epaper

## Acknowledgments

SS and THR acknowledge the support from the Australian Research Council under Linkage Scheme (LP160100450). THR acknowledges the support from the Australian research Council under the DECRA Scheme (DE170101346).

## Author contributions

THR and FV conceived and supervised the project. SS and THR 5 developed the mathematical model. SS generated the results for the manuscript. AA and FV developed the online platform. THR and FV wrote the manuscript. All authors reviewed and approved the final manuscript.

## Data and materials availability

All data is available in the main text or the supplementary 10 materials.

